# Comparative of the Effectiveness of Different Objective Method for Infraclavicular Brachial Plexus Block Success; Which One of Them is A Reliable and Early Indicator?

**DOI:** 10.1101/2021.06.26.21259567

**Authors:** Abdulhakim Şengel, Mahmut Alp Karahan, Nuray Altay, Orhan Binici, Veli Fahri Pehlivan, Ahmet Atlas

## Abstract

**BACKGROUND:** Traditional methods that evaluate the success of peripheral nerve block have been replaced by methods that allow objective evaluations over time. Multiple objective techniques for peripheral nerve block have been discussed in the literature.

**OBJECTIVE:** This study aims to investigate whether perfusion index (PI), non-invasive tissue hemoglobin monitoring (SpHb), tissue oxygen saturation (StO_2_), tissue hemoglobin index (THI) and body temperature are reliable and objective methods to evaluate the adequacy of infraclavicular blockage.

**DESIGN:** A prospective observational study.

**SETTING:** Single center, Department of Anesthesiology, Harran University Medical Faculty, Turkey, from February 2019 to December 2019.

**PATIENTS:** A total of 100 patients scheduled to undergo forearm surgery.

**INTERVENTION:** Ultrasound-guided infraclavicular block (ICB) in 100 patients undergoing forearm surgery.

**MAIN OUTCOME MEASURES:** PI, SpHb, StO_2_, THI and Body Temperature measurements were recorded 5 min before the block procedure, right after the procedure, and until the 25th minute after the procedure at 5-min intervals. These values were compared between the blocked limbs and non-blocked limbs while being statistically compared between the successful and failed block groups.

**RESULTS:** Although there were significant differences between the groups of blocked extremity and non-blocked extremity in terms of StO_2_ (P = 0.001), THI (P = 0.001), PI (P = 0.001) and body temperature (P = 0.001), there was no significant difference between these groups in terms of SpHb (P > 0.05). Moreover, a significant difference was detected between the groups of successful block and failed block in terms of StO_2_ (P = 0.002), PI (P = 0.002) and body temperature (P = 0.005), while there was no significant difference between these groups in terms of THI (P > 0.05) and SpHb (P > 0.05).

**CONCLUSION:** StO_2_, PI and body temperature measurements are the simple, objective, noninvasive techniques to be used to evaluate success of block procedures. According to The receiver operating characteristic (ROC) analysis, StO_2_ is the specific parameter with the highest sensitivity among these parameters.

**TRIAL REGISTRATION:** ACTRN12621000588897

## INTRODUCTION

At present, peripheral nerve blocks are increasingly used for both anesthesia and postoperative analgesia, particularly in extremity surgeries.^1^ These blocks are the most common regional anesthesia method after spinal anesthesia. Brachial plexus block, which is one of the most used peripheral nerve blocks, allows regional block of the shoulder, arm, wrist and forearm.^2^ Infraclavicular block is a brachial plexus block technique that is easy to apply under ultrasonography (USG) guidance in upper extremity surgeries and can be used in outpatient surgeries.^3^ Whether the peripheral nerve block for anesthesia has been successful is determined by evaluating the levels of sympathetic, sensory and motor blocks.^4^

Many different techniques have been employed throughout the development process of peripheral nerve blocking depending on whether a block procedure was concluded to be successful or not. The comments on this subject were based on mostly traditional methods and caused certain difficulties on a subjective basis, as well as different interpretations of assessment varying on a personal basis. However, the necessity of quality communication with the patient^5^ led researchers to look for new ways and required non-invasive methods that do not require patient cooperation and provide faster evaluation. Therefore, traditional methods have been replaced by the methods that allow objective evaluations over time.

These objective methods were introduced based on the evaluation of the local vasodilation, increased blood flow and increased skin temperature resulting from the sympathetic blockage in the blocked area after the block procedure.^5^

Multiple objective techniques for peripheral nerve block have been discussed in the literature. These techniques include methods such as perfusion index (PI), plethysmographic variability index (PVI), non-invasive tissue hemoglobin monitoring (SpHb), tissue oxygen saturation (StO_2_), tissue hemoglobin index (THI), and body temperature.^6-13^

This study is intended to compare the superiority of near infrared spectroscopy (NIRS), StO_2_, THI, PI, SpHb, and body temperature methods along with the routine vital signs and traditional methods in assessing the block success and adequacy of the infraclavicular brachial plexus block procedures that we applied under USG guidance in cases undergoing forearm surgery.

## METHODS

### Ethics

Ethical approval for this study was provided by Harran University Medical Faculty’s Ethics Committee, Şanlıurfa, Turkey (Chairperson Prof Dr. Fuat Dilmeç) on 14 February 2019. It was registered with the number 377625 at the clinical research center of Australian New Zealand Clinical Trials Registry (ANZCTR). Our study was performed between February 2019 and December 2019.

### Patient Selection

We calculated the sample size according to the results of with previous similar studies and the first 15 patients in the study. From these differences and assuming a two-tailed α value of 0.05 (sensitivity 95%) and a β value of 0.20 (study power: 80%, effect size: 0.80), we determined that at least 38 patients were required for our study by statistical software Package G Power (version 3.1.9.2; Franz Faul & Edgar Erdfelder, Trier, Germany). We decided to enroll 100 ASA I-II patients aged 18-65 who will undergo forearm surgery. The patients who were accepted to the study were informed about the study both verbally and in writing. Moreover, informed consent was issued for patients who accepted to participate in the study.

Exclusion criteria included patients who did not want to participate in the study; patients who were not included in ASA-I and ASA-II; patients who were not in the age range of 18–65; presence of a previous neurological sequel in the limb to be blocked; patients contraindicated for infraclavicular block because of the presence of open wound, infection, and coagulopathy disorder in the area to be blocked; patients with a history of allergy to amide group Local Anesthetics (LA); patients who could not cooperate; patients with kidney and liver failure, pregnant and breastfeeding patients; and patients with contralateral phrenic paralysis.

### Procedures Infraclavicular Brachial Plexus

Block Patients undergoing forearm surgery were taken to the preoperative preparation unit at 23-24°C before surgery. Standard monitoring, including non-invasive arterial blood pressure, electrocardiography (ECG), and peripheral oxygen saturation (SpO2), was performed in the patients who were taken to the operating table. In the upper extremity where no operation was scheduled, after the peripheral vascular access was established with an 18- or 20-gauge cannula, an infusion of 10 ml/kg of 0.9% NaCl liquid was started for maintenance purposes. Midazolam (Zolamid 5 mg/mL) of 0.02-0.03 mg/kg was administered i.v. to the patients for sedative purposes. By communicating with the patients, 2 L/min of oxygen was given to the patients via the nasal route during the block procedure and during the follow-up. For recording StO_2_ and THI measurements, NIRS probe (Inspectra(tm) StO_2_ Spot Check - Model 300 (Hutchinson Technology Inc., Hutchinson, MN, USA) device probe) was placed on the palm of the hand to cover the thenar ridge of the hand to be blocked as well as on the palm of the other hand. A special pulse oximetry sensor (R1 25 SpHb SpO2 SpMet Adult Pulse CO-Oximeter Adhesive Sensor) was placed on the middle fingers of both hands.

This sensor (Masimo IC model: the RDS7A device (USA)) was used to obtain the data to be recorded. Moreover, 5 min before and during the procedure, systolic blood pressure (SBP) and diastolic blood pressure (DBP), heart rate (HR) and SpO2, as well as the StO_2_ and THI data obtained from the NIRS device, and the PI and SpHb data from in the Pulse CO-Oximeter device were all recorded. After these basal values were recorded, the procedure stage was started.

To perform the block, it was planned to move towards the axillary artery with USG guidance using a 22 gauge, 50 mm, insulated facet tip needle (B. Braun Stimuplex, Melsungen AG, Germany). Before the intervention with the block needle, 2 mL lidocaine was administered for skin-subcutaneous infiltration to the area planned to be injected. After the skin infiltration, the long axis method was employed and the lateral, medial and posterior cords around the axillary artery were accessed with simultaneous USG-imaging, and the block needle was directed to the nearest of each cord with local anesthetic. After reaching the desired area, after negative aspiration test (by repeating this test after every 5 mL LA injection), the prepared local anesthetic solution was injected into the described areas. To avoid an unwanted situation, when the distribution of LA could not be completely confirmed on the US screen, when any resistance was encountered at the time of injection, or when the patient described pain during the injection, the needle was redirected with fine maneuvers. Local anesthetic solution of 15 ml of 2% lidocaine (Lidon 100 mg/5ml On Farma), and 15 ml of 0.5% bupivacaine (Buvasin 5 mg/ml, 20 ml, Vem İlaç, Turkey) were prepared. After the entire solution was administered into the desired areas, the U-shaped distribution of the solution around the axillary artery was observed using USG.

### Clinical Evaluation Block Assessment

The anesthetist performing the block operation and the anesthetist performing the post-procedure follow-up consisted of different people. For follow-up, the minute 0 was accepted as the moment when the procedure was terminated and the needle was removed from the skin. Block procedure was applied to 100 patients by the same anesthesiologist. The recording of the data for monitoring was performed by a different anesthesiologist. After the block procedure (the moment of removal of the needle from the skin was accepted as the minute 0), HR, SBP, DBP, and SpO2 values were recorded at the minutes 0, 5, 10, 15, 20, and 25. Moreover, the StO_2_, THI, PI, SpHb and body temperature (the temperature of the extremities was measured with an infrared thermometer (Mesilife DT-8806) from the intermediate region of the two arteries, not corresponding to the radial and ulnar artery traces on the inner side of both wrists) data, was recorded from both the blocked extremity and the other extremity one by one at the minutes 0, 5, 10, 15, 20, and 25.

Furthermore, 30 minutes after the block procedure, it was recorded whether the block was holding or not after the evaluation with Bromage scale, the sensory test for loss of cold sensation and Pin-prick test in the extremity where the block was performed.

### Statistical Method

Compliance of numerical data to normal distribution was tested with Shapiro–Wilk test. Mann– Whitney U test was used to compare non-normally distributed variables between the groups of successful and failed block. Furthermore, the Wilcoxon test was used to compare the non-normally distributed variables between the block group and the control group, while the Freidman test was used to compare the measurements obtained at seven different times (intragroup comparison), and Dunn’s multiple comparison test was used to determine the times reported to be significant. Inter-method consistency was tested with Kappa statistics. Mean ± standard deviation values were given for summarizing numerical variables, and numbers and % values were given for categorical variables. SPSS windows 24 version was used for statistical analysis, and analysis results were evaluated at 95% confidence interval with a p value of<0.05 considered to be statistically significant. ROC analysis was used between the block and control groups to determine the sensitivity and specificity of the parameters found to be significant in the evaluation of the block procedure, and the areas under the curve of each predictor were calculated and compared with each other.

## RESULTS

A total of 100 patients aged 18–65 years who underwent forearm surgery under infraclavicular plexus block were included in the study. The mean age was 37.37±13.54 years. Table 1 shows the demographic data of the patients. Block failure occurred in 5 patients (5%) who were then switched to general anesthesia. There were no complications related to the technique or local anesthetic injection.

**Table 1.** Demographic data of the cases

| Variables | | Descriptive statistics (n = 100) Mean±SD | |
| --- | --- | --- | --- |
| Age | | $37.37 \pm 13.54$ (min = 18-max = 64) | |
| Height | | $168.74 \pm 7.04$ | |
| Weight | | $74.92 \pm 12.94$ | |
| BMI | | $26.39 \pm 4.78$ | |
|  |  | Number | % |
| Gender | Male | 54 | 54,0 |
|  | Female | 46 | 46,0 |
| ASA | 1.00 | 45 | 45,0 |
|  | 2.00 | 55 | 55,0 |
| Additional disease | No | 74 | 74,0 |
|  | Yes | 26 | 26,0 |
| Smoking | No | 66 | 66,0 |
|  | Yes | 34 | 34,0 |

BMI: Body Mass Index, ASA: American Society of Anesthesiology Risk Classification

Compared to the pre-procedure period, the mean percentage change of HR variable decreased by 2% at the minutes 0, 5, and 10, 4% at the minute 15, and 3% at the minutes 20 and 25. A significant difference was observed in HR values starting at the 10th minute, compared to the pre-procedure values (P = 0.001). Compared to its pre-procedure value, the mean percentage change of SBP presented no change at the minute 0 but decreased by 2% at the 5th, 10th and 20th minutes, 3% at the 15th minute and 1% at the 20th minute. Compared to its pre-procedure value, the mean percentage change of DBP, however, increased by 1% at the minutes 0 and 5, 2% at the 10^th^, 15^th^ and 20^th^ minutes, and 3% at the 25^th^ minute. The change in the mean arterial pressure (MAP) over time was not found significant (P = 0.391). The change in SpO2 over time was not significant either (p > 0.05).

When we examine the differences of temperature, StO_2_, THI, SpHb and PI values between the groups, after the minute 0, all measurements of temperature (P = 0.001), StO_2_ (P = 0.001), THI (P = 0.001) and PI (P = 0.001) but SpHb revealed significant differences between the blocked limb and the non-blocked limb (Table 2).

**Table 2.** Comparison of the measurements between the blocked limb and the non-blocked limb

| Time | Variables | Blocked limb (n: 100) | Non-blocked limb (n: 100) | Mean difference [95% CI] | P |
| --- | --- | --- | --- | --- | --- |
| Pre-procedure | Temperature | 35.81 ± 0.81 | 35.78 ± 0.77 | 0.04 [-0.02 -1.39] | 0,169 |
|  | StO<sub>2</sub> | 81.33 ± 5.89 | 81.33 ± 5.72 | -0.15 [-0.91 --0.39] | 0.532 |
|  | THI | 12.61 ± 1.95 | 12.4 ± 2.14 | 0.17 [-0.25 --0.39] | 0.265 |
|  | PI | 3.38 ± 2.12 | 3.61 ± 2.62 | -0.25 [-0.61 --0.39] | 0,153 |
|  | SpHb | 11.69 ± 0.98 | 11.74 ± 0.98 | -0.03 [-0.14 --0.39] | 0.743 |
| Minute 0 | Temperature | 35.9 ± 0.76 | 35.74 ± 0.79 | 0.15 [-0.09 -5.33] | **0.001 \*** |
|  | StO<sub>2</sub> | 83.01 ± 5.76 | 80.18 ± 5.8 | 2.5 [1.59 -5.44] | **0.001 \*** |
|  | THI | 12.91 ± 2.15 | 12.25 ± 1.8 | 0.61 [0.18 -5.44] | **0.001 \*** |
|  | PI | 4,06 ± 2,33 | 3.46 ± 2.57 | 0.55 [0.11 -5.44] | **0.001 \*** |
|  | SpHb | 11.74 ± 1 | 11.72 ± 0.98 | 0.04 [-0.08 -5.44] | 0,686 |
| Minute 5 | Temperature | 36.15 ± 0.64 | 35.73 ± 0.81 | 0.4 [-0.32 -9.86] | **0.001 \*** |
|  | StO<sub>2</sub> | 85.8 ± 5.43 | 79.7 ± 5.91 | 5.77 [4.83 -12.14] | **0.001 \*** |
|  | THI | 14,09 ± 2,77 | 12.26 ± 2.12 | 1.73 [1.16 -12.14] | **0.001 \*** |
|  | PI | 5,92 ± 2,57 | 3.21 ± 2.39 | 2.57 [2.02 -12.14] | **0.001 \*** |
|  | SpHb | 11.79 ± 0.94 | 11.77 ± 1.02 | 0.04 [-0.1 -12.14] | 0,670 |
| Minute 10 | Temperature | 36.35 ± 0.58 | 35.63 ± 0.95 | 0.7 [0.53 -8.21] | **0.001 \*** |
|  | StO<sub>2</sub> | 87.09 ± 4.97 | 79.34 ± 6.04 | 7.31 [6.33 -14.85] | **0.001 \*** |
|  | THI | 14.59 ± 2.96 | 12.26 ± 2.16 | 2.22 [1.65 -14.85] | **0.001 \*** |
|  | PI | 6.67 ± 2.47 | 3.14 ± 2.3 | 3.41 [2.81 -14.85] | **0.001 \*** |
|  | SpHb | 11.78 ± 0.99 | 11.75 ± 1 | 0.05 [-0.1 -14.85] | 0,629 |
| Minute 15 | Temperature | 36.5 ± 0.54 | 35.68 ± 0.81 | 0.8 [0.69 -15.05] | **0.001 \*** |
|  | StO<sub>2</sub> | 87.48 ± 4.97 | 79.05 ± 6.16 | 8.02 [6.92 -14.45] | **0.001 \*** |
|  | THI | 14.4 ± 2.88 | 12.15 ± 1.91 | 2.13 [1.6 -14.45] | **0.001 \*** |
|  | PI | 6.52 ± 2.42 | 2.98 ± 2.16 | 3.44 [2.89 -14.45] | **0.001 \*** |
|  | SpHb | 11.7 ± 0.94 | 11.74 ± 1.03 | -0.03 [-0.17 -14.45] | 0.852 |
| Minute 20 | Temperature | 36.6 ± 0.52 | 35.68 ± 0.8 | 0.89 [0.78 -15.93] | **0.001 \*** |
|  | StO<sub>2</sub> | 87.48 ± 4.54 | 78.86 ± 6.17 | 8.26 [7.19 -15.28] | **0.001 \*** |
|  | THI | 14.2 ± 2.65 | 12.12 ± 2.23 | 1.99 [1.47 -15.28] | **0.001 \*** |
|  | PI | 6.43 ± 2.51 | 2.75 ± 1.94 | 3.55 [3.02 -15.28] | **0.001 \*** |
|  | SpHb | 11.7 ± 1.01 | 11.73 ± 1.03 | -0.01 [-0.17 -15.28] | 0.934 |
| Minute 25 | Temperature | 36.69 ± 0.51 | 35.66 ± 0.81 | 0.99 [0.87 -16.77] | **0.001 \*** |
|  | StO<sub>2</sub> | 88.01 ± 4.36 | 78.55 ± 6.06 | 9.05 [8.02 -17.42] | **0.001 \*** |
|  | THI | 13.88 ± 2.69 | 11.96 ± 2.27 | 1.88 [1.4 -17.42] | **0.001 \*** |
|  | PI | 6.35 ± 2.42 | 2.75 ± 1.94 | 3.55 [3.04 -17.42] | **0.001 \*** |
|  | SpHb | 11.7 ± 0.99 | 11.71 ± 1.02 | 0.02 [-0.14 -17.42] | 0.763 |

**StO<sub>2</sub>:** Tissue Oxygen Saturation, **THI:** Tissue Hemoglobin Index, **PI:** Perfusion Index, **SpHb:** Non-invasive Total Hemoglobin

When the groups of successful block and failed block were compared, StO_2_ was reported to be significantly higher in all measurements starting from the 5th minute after the block (P = 0.002-0.011). The temperature was reported to be highly significant in other measurements starting from the 15th minute (P = 0.044-0.012). PI was reported to be significant in other measurements starting from the 5th minute but not at the 15th minute (P = 0.044-0.002). There was no significant difference between the groups in terms of SpHb value (Table 3).

**Table 3.** Comparison of the measurements between the groups of successful block and failed block

| Time | Variables | Successful Block (n = 95) | Failed Block (n = 5) | Mean difference [95% CI] | P |
| --- | --- | --- | --- | --- | --- |
| Pre-procedure | Temperature | 35.81 ± 0.82 | 35.8 ± 0.67 | 0.01 [-0.73 -0.76] | 0.606 |
|  | StO<sub>2</sub> | 81.33 ± 5.89 | 78.4 ± 6.31 | 2.93 [-2.45 -8.3] | 0.401 |
|  | THI | 12.61 ± 1.95 | 11.9 ± 2.15 | 0.71 [-1.08 -2.49] | 0,613 |
|  | PI | 3.38 ± 2.12 | 3.08 ± 2.43 | 0.3 [-1.64 -2.24] | 0.722 |
|  | SpHb | 11.69 ± 0.98 | 12.12 ± 0.45 | -0.43 [-1.31 -0.45] | 0,181 |
| Minute 0 | Temperature | 35.9 ± 0.76 | 35.78 ± 0.63 | 0.12 [-0.57 -0.81] | 0,442 |
|  | StO<sub>2</sub> | 83.01 ± 5.76 | 76.4 ± 7.09 | 6.61 [1.31 -11.91] | 0.052 |
|  | THI | 12.91 ± 2.15 | 11.9 ± 2.13 | 1.01 [-0.94 -2.97] | 0,275 |
|  | PI | 4,06 ± 2,33 | 3.24 ± 2.93 | 0.82 [-1.33 -2.97] | 0.355 |
|  | SpHb | 11.74 ± 1 | 12.14 ± 0.45 | -0.4 [-1.29 -0.5] | 0,203 |
| Minute 5 | Temperature | 36.15 ± 0.64 | 35.78 ± 0.65 | 0.37 [-0.22 -0.95] | 0,151 |
|  | **StO<sub>2</sub>** | **85.8 ± 5.43** | **79.2 ± 4.87** | **6.6 [1.67 -11.53]** | **0.011** |
|  | THI | 14,09 ± 2,77 | 12.16 ± 2.23 | 1.93 [-0.57 -4.43] | 0.090 |
|  | **PI** | **5,92 ± 2,57** | **3.03 ± 2.49** | **2.89 [0.56 -5.23]** | **0.021** |
|  | SpHb | 11.79 ± 0.94 | 12.02 ± 0.64 | -0.23 [-1.07 -0.62] | 0.420 |
| Minute 10 | Temperature | 36.35 ± 0.58 | 35.84 ± 0.73 | 0.51 [-0.02 -1.05] | 0.097 |
|  | **StO<sub>2</sub>** | **87.09 ± 4.97** | **78.2 ± 6.34** | **8.89 [4.32 -13.47]** | **0.005** |
|  | THI | 14.59 ± 2.96 | 12.46 ± 2.68 | 2.13 [-0.56 -4.81] | 0,110 |
|  | **PI** | **6.67 ± 2.47** | **4.34 ± 2.89** | **2.33 [0.07 -4.59]** | **0.044** |
|  | SpHb | 11.78 ± 0.99 | 12.14 ± 0.52 | -0.36 [-1.25 -0.52] | 0,268 |
| Minute 15 | **Temperature** | **36.5 ± 0.54** | **35.9 ± 0.68** | **0.6 [0.11 -1.1]** | **0.044** |
|  | **StO<sub>2</sub>** | **87.48 ± 4.97** | **79.2 ± 4.44** | **8.28 [3.77 -12.79]** | **0.003** |
|  | THI | 14.4 ± 2.88 | 12 ± 2.57 | 2.4 [-0.21 -5.01] | 0.080 |
|  | PI | 6.52 ± 2.42 | 4.52 ± 2.8 | 2 [-0.22 -4.22] | 0.053 |
|  | SpHb | 11.7 ± 0.94 | 12.1 ± 0.6 | -0.4 [-1.25 -0.45] | 0,282 |
| Minute 20 | **Temperature** | **36.6 ± 0.52** | **35.9 ± 0.69** | **0.7 [0.22 -1.18]** | **0.019** |
|  | **StO<sub>2</sub>** | **87.48 ± 4.54** | **80.2 ± 5.26** | **7.28 [3.12 -11.45]** | **0.007** |
|  | THI | 14.2 ± 2.65 | 12.3 ± 2.75 | 1.9 [-0.52 -4.32] | 0,141 |
|  | **PI** | **6.43 ± 2.51** | **3.78 ± 1.53** | **2.65 [0.4 -4.91]** | **0.012** |
|  | SpHb | 11.7 ± 1.01 | 12.18 ± 0.54 | -0.48 [-1.39 -0.42] | 0,184 |
| Minute 25 | **Temperature** | **36.69 ± 0.51** | **35.98 ± 0.62** | **0.71 [0.24 -1.17]** | **0.012** |
|  | **StO<sub>2</sub>** | **88.01 ± 4.36** | **79.8 ± 5.22** | **8.21 [4.21 -12.22]** | **0.002** |
|  | THI | 13.88 ± 2.69 | 12.98 ± 2.7 | 0.9 [-1.55 -3.35] | 0,389 |
|  | **PI** | **6.35 ± 2.42** | **3.36 ± 1.05** | **2.99 [0.83 -5.16]** | **0.002** |
|  | SpHb | 11.7 ± 0.99 | 12.3 ± 0.44 | -0.6 [-1.49 -0.29] | 0.093 |

**StO<sub>2</sub>:** Tissue Oxygen Saturation, **THI:** Tissue Hemoglobin Index, **PI:** Perfusion Index, **SpHb:**

In the ROC analysis of the variables, the most significant result in terms of the values of StO_2_ was found to be at the fifth minute, the Cutoff value to be 78.5, the Area under the Curve to be 0.806, the sensitivity to be 93% and specificity to be 58% (Fig. 1). The most significant result of PI is at the 25th minute with the Cutoff value being 2.45, the Area under the Curve to be 0.791, the sensitivity to be 89% and the specificity to be 51% (Fig. 2). The ROC analysis result where temperature was the most significant was at the 15th minute with the Area under the Curve being 0.833, the Cutoff value being 36.45, sensitivity being 63%, and specificity being 60% (Fig. 3).

**Figure 1:**
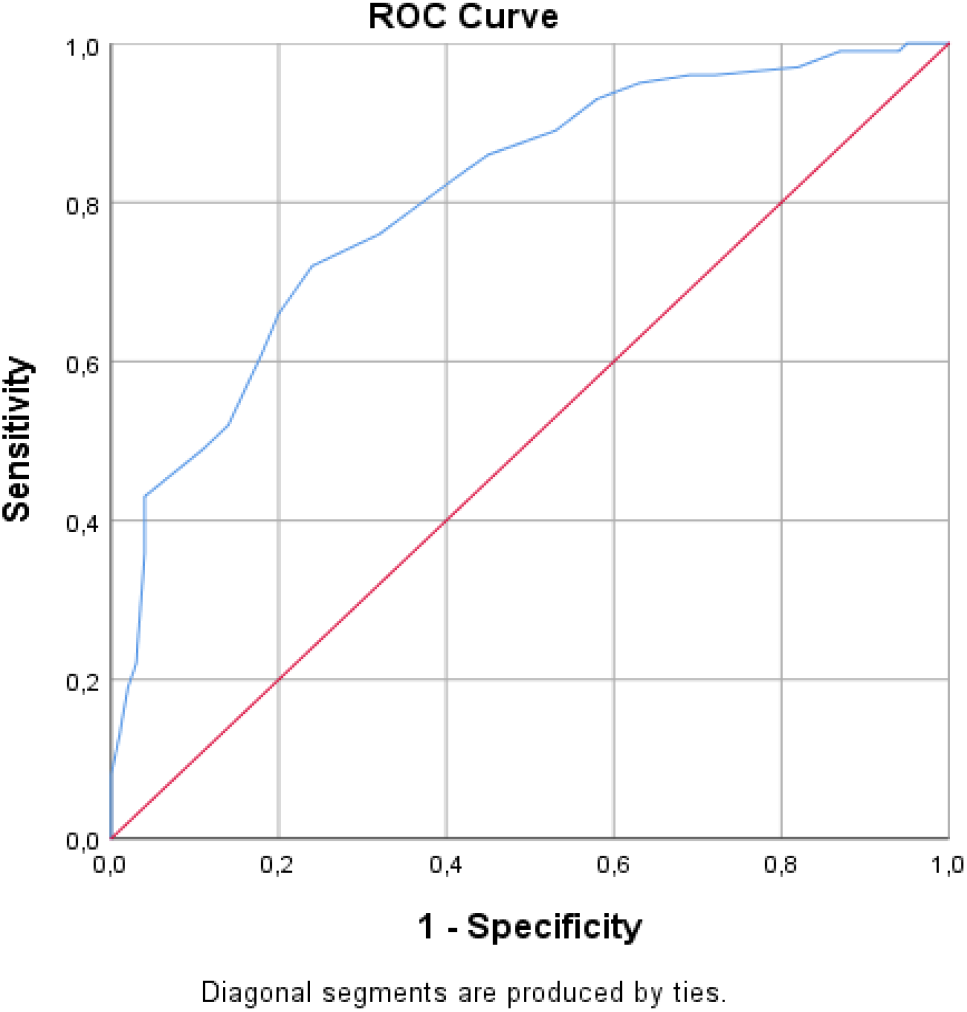
The ROC Analysis of StO_2_ between the Block Group and the Control Group is the ROC curve of StO_2_ for the STO value. Cutoff value: 78.5, sensitivity: 93%, and specificity: 58% (Fig. 1).

**Figure 2:**
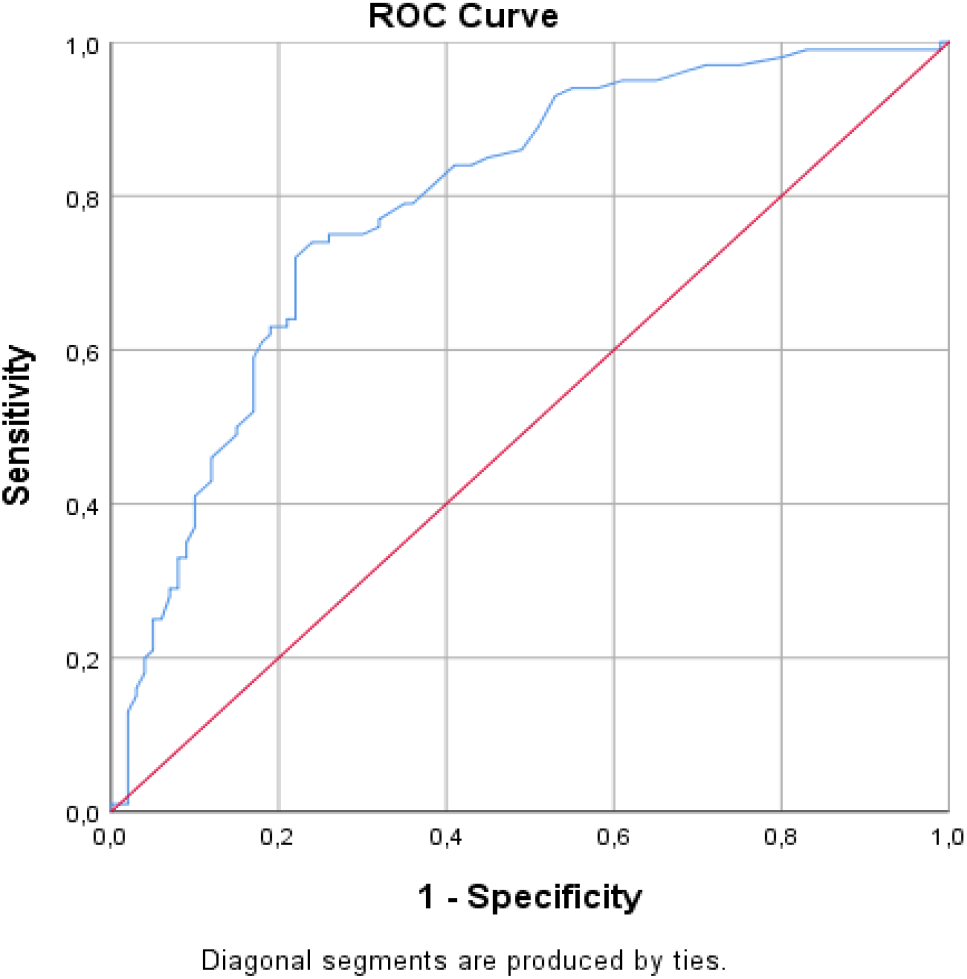
The ROC Analysis of PI between the Block Group and the Control Group is the ROC curve for PI value. Cutoff value: 3.15, sensitivity: 82%, and specificity: 39% (Fig. 2).

**Figure 3:**
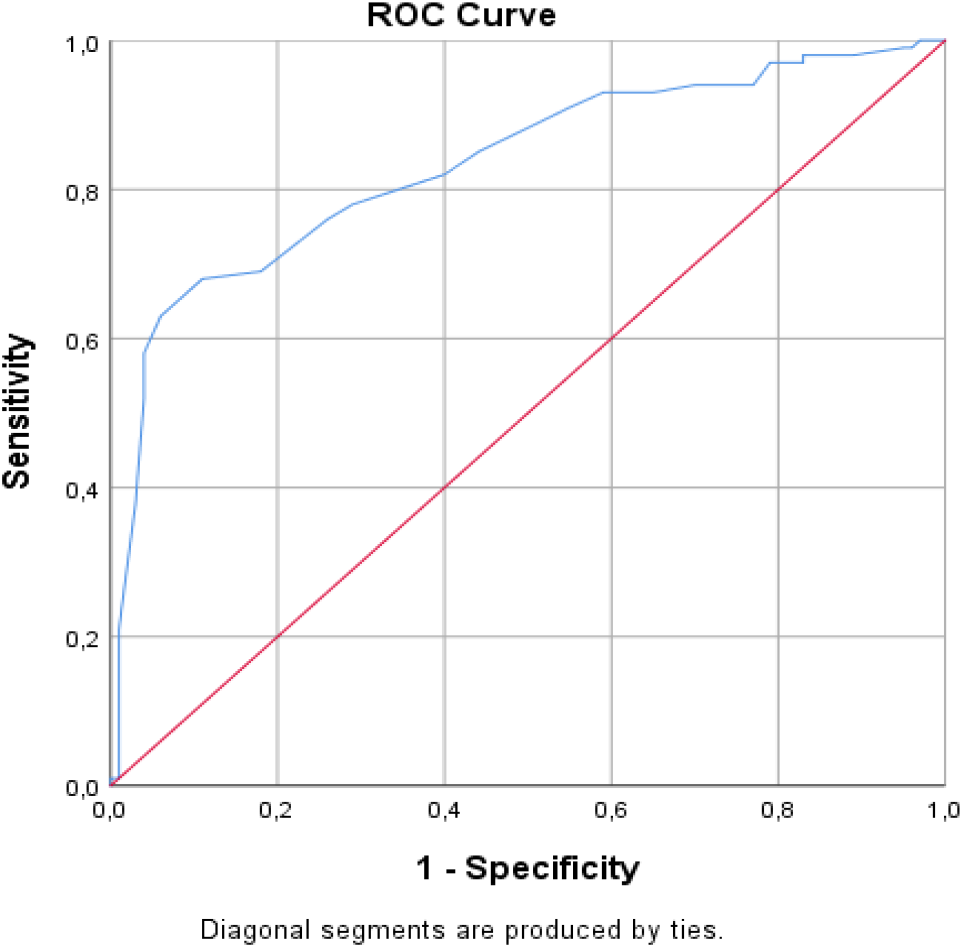
The ROC Analysis of temperature between the Block Group and the Control Group is the ROC curve for the temperature value. Cutoff value: 36.45, sensitivity: 63%, and specificity: 60% (Fig. 3).

Compared to the pre-procedure time, the mean percentage change of PI increased by 29% at minute 0, 110% at the 5th minute, 150% at the 10th minute, 146% at the 15th minute, 144% at the 20th minute, and 138% at the 25th minute. Compared to the pre-procedure time, the mean percentage change of StO_2_ increased by 2% at minute 0, 5% at the 5th minute, 7% at the 10th and 15th minutes, and 7% at the 20th and 25th minutes. Compared to the pre-procedure time, the mean percentage change of THI variable increased by 3% at the minute 0, 12% at the 5th minute, 16% at the 10th minute, 14% at the 15th minute, 13% at the 20th minute and 11% at the 25th minute. Compared to pre-procedure time, the mean percentage change of the temperature variable presented no change at minute 0 and increased by 1% at the 5th and 10th minutes and by 2% at the 15th, 20th and 25th minutes. Compared to pre-procedure time, the mean percentage change of the SpHb variable presented no change at the minutes 0, 15, 20 and 25 but increased by 1% at the 5th and 10th minutes (Fig. 4).

**Figure 4:**
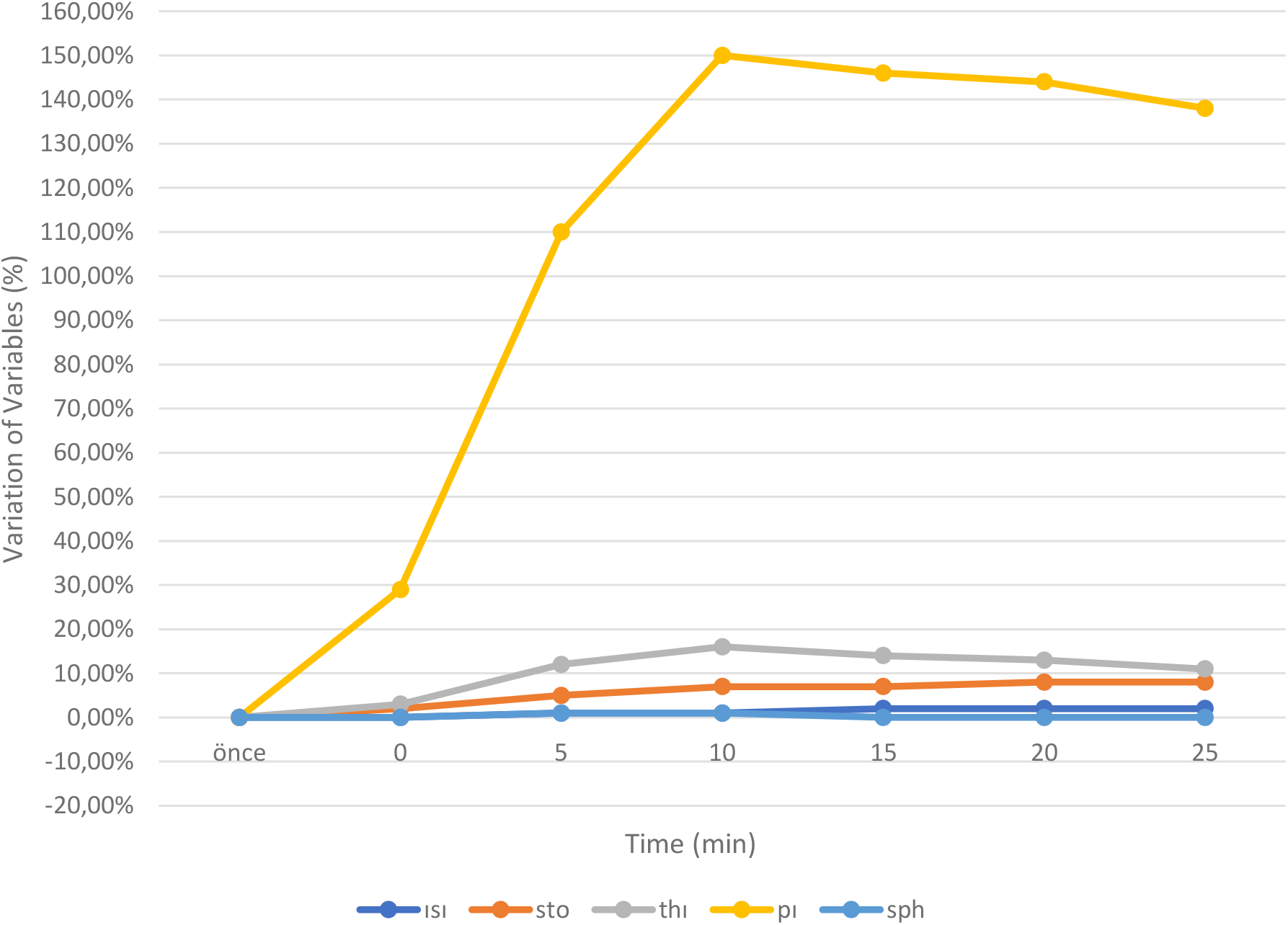
The percentage changes in mean values of the variables of temperature, StO_2_, THI, PI, SpHb

When we evaluated the success of the block with traditional methods before sending the 100 patients undergoing the block procedure to the operating room, no sensory loss was observed in 5 patients in the Pinprick test and the sensory test for loss of cold sensation. In 77 patients, it was observed that there was no feeling of tactile sensation and pain. A complete consistency was observed between the Pinprick test and the sensory test for loss of cold sensation (Kappa = 1.000, P = 0.001) (Table 4).

**Table 4.** Confirmation of block success and block failure with traditional diagnostic methods

|  |  | Number | % |
| --- | --- | --- | --- |
| Bromage Scale | motor strength is reduced but arm is moving | 13 | 13,0 |
|  | arm is immobile but fingers are moving | 41 | 41,0 |
|  | full block no movement in hand and arm | 46 | 46,0 |
| Pin-prick test | no sensory block | 5 | 5,0 |
|  | There is tactile sensation and no pain | 18 | 18,0 |
|  | There is no tactile sensation and pain | 77 | 77,0 |
| Sensory test for loss of cold sensation | no sensory block | 5 | 5,0 |
|  | analgesia (the patient feels the touch but does not have the cold sensation) | 18 | 18,0 |
|  | anesthesia (patient does not feel the touch) | 77 | 77,0 |

## DISCUSSION

The current study is intended to investigate whether noninvasive, objective and observer-independent methods of PI, SpHb, StO_2_, THI and Body Temperature are an early and reliable predictor of block success or failure in patients undergoing forearm surgery. Our study is the study with the biggest sample size and the first study in which objective methods that are normally used separately to measure block success are evaluated all together. Temperature, StO_2_, THI and PI values were significantly different in all these parameters -except for SpHb-between the block group and the control group after minute 0. We determined PI and StO2 as objective methods since they started at the earliest from the 5th minute and continued in the late period.

NIRS is a non-invasive method used to detect StO_2_ based on the microcirculation perfusion in the area of measurement where near infrared light of different wavelengths (680–800 nm) is used for measurement. NIRS detects differences in oxygen saturation caused by changes in tissue oxygen consumption or oxygen delivery. It is mostly used as a cerebral oximeter monitor in critically severe cases.^14^ Although there have been publications about the use of StO_2_ in evaluating peripheral nerve block success, there are few studies in the literature regarding its use to measure the success of peripheral block.^9,15-18^

In the first study, which showed that NIRS can capture the increase in local tissue perfusion occurring after the block, a 15% increase in oxygen saturation was detected in the limb that was performed block procedure after the brachial plexus blockage (interscalene, supraclavicular, infraclavicular or axillary). Significant increases in StO_2_ values were observed within 5 min after the completion of the block, which occurred before the patients perceived sensory and motor loss.^8^ Various other subsequent studies in which peripheral nerve blocks were performed in the lower and upper extremities reported that there were statistically significant differences in StO_2_ values in the blocked limbs and the control limbs.^15^ In these studies, the increases were significant after the 5th minute, and there was a significant difference between the blocked limb and the non-blocked limb and that StO_2_ could successfully demonstrate peripheral nerve block as a noninvasive parameter. Our study has a greater sample size compared to other studies and was conducted on 100 cases. Our results are similar to the previous results; in the comparison between the groups of successful and failed block, there is a significant increase starting from the 5th minute and including the 25th minute.

Another parameter other than StO_2_, which we could measure with the NIRS method, was THI. THI is the tissue hemoglobin concentration used for StO_2_ measurement and measured quantitatively from the thenar ridge using a probe. It is also accepted as a measurement of hemoglobin signal strength, which is useful for determining whether the StO_2_ sensor is best positioned on the muscle.^10^ In a single study related to its use in peripheral block, Karahan et al. investigated the change in tissue oxygen saturation in patients who underwent infraclavicular block and studied THI values along with StO_2_ and reported that THI was a sign of vasodilation and showed a significant difference over time in the blocked limb.^9^ In our study, although we reported a significant difference in THI values between the blocked extremity and control extremity groups, no significant difference was reported between the groups of successful and failed block.

Another objective value by which the brachial plexus block success is followed in the literature is PI. The PI is a numerical value that reflects the relative strengths of the different components of the infrared signal returning from the monitoring area, showing the ratio of pulsatile blood flow to non-pulsatile or static blood in the peripheral tissue. It has been shown to reflect real-time changes in peripheral blood flow based on pulse oximetry continuously and noninvasively. Since PI is an indicator of peripheral blood flow and peripheral blood flow in the upper extremities is a clinical sign of an effective block, increased PI on the side of the block can be a reliable approach to determine the effectiveness of infraclavicular block.^5^

It has been shown that sympathetic blockage is correctly detected by evaluating the increase in PI shortly after epidural block, pediatric caudal block, stellate ganglion block and interscalene nerve block.^11,12,19,20^ The study by Abdelnasser et al. on the use of perfusion index measured by the pulse oximetry method in predicting the success of supraclavicular brachial plexus block concluded that high PI values had 100% sensitivity and specificity for successful block.^21^ Separate studies by Kuş et al. and Nieuwveld et al. concluded that PI is a successful infraclavicular indicator in patients in whom infraclavicular block was applied.^5,22^ Similar to the studies published in the literature, based on the comparison of the PI value between the groups, our study reported a significant difference between the blocked and non-blocked groups starting from the minute 0. The data we obtained in our study were similar to the data obtained from previous studies. When we examine the significant difference between the groups of successful block and failed block, a significant difference emerged at the 5th minute and afterwards in terms of PI.

Another popular objective parameter investigated is skin temperature. It is claimed tissue blood flow increases because of the vasodilation resulting from the sympathetic block in such blocks and increase in the skin temperature, and this increase in skin temperature shows block success with higher specificity and sensitivity compared to the pinprick and cold sensation tests.^23^ With this theory, studies investigated whether a simple infrared thermometer could reliably predict block efficacy after infraclavicular brachial plexus block and reported that the measurement of skin temperature with infrared thermometry would be an inexpensive and easy-to-use predictor of the early stage of block success or failure in a particular nerve distribution.^7^

In a study evaluating distal skin temperature to predict the success of lateral infraclavicular block, the authors concluded that the temperature difference predicts a successful block with 92% sensitivity (95% confidence interval (CI), 83% -97%). They determined the positive predictive value of the test as 95% (CI, 86-98%). Consequently, they reported that the difference in skin temperature measured between the blocked and non-blocked hands is a valid and reliable diagnostic test to predict successful lateral infraclavicular block.^24^ In the study where thermochromic nail polish was used to predict the success of infraclavicular brachial plexus block, they reported a positive predictive value of 96% and a sensitivity of 94% for thermochromic nail color change. Consequently, this study reported that thermochromic nail polish color change is a valid and reliable indicator for predicting block success.^25^

In our study, when we measured the temperature change between the blocked limb and the non-blocked limb with an easy-to-use infrared thermometer device, a significant difference was observed between the blocked and non-blocked groups starting from the minute 0. However, when we examine the difference between the groups of successful block and failed block, no significant difference was observed in the first 10 minutes while a significant increase was found in the group of successful block after the 15th minute. It was also observed that this increase continued at the 20th and 25th minutes as well.

Developing technology leads scientists to look for alternatives. Considering that the technology is noninvasive for these reasons and its ability to provide continuous, real-time data can be associated with events occurring at the bedside, SpHb monitoring offers us a new paradigm, new possibilities for better and healthier follow-up.^26^

The study where Bergek et al. measured noninvasive hemoglobin and Pleth Variability Index after brachial plexus block, it was evaluated that SpHb values increased compared to basal value and this was a significant difference. It was stated that there was no significant difference in this parameter in the control arm. In the same study, it was stated that SpHb was a measure of intravascular Hb concentration, and it was stated that vascular structures were affected depending on the anesthesia technique applied, and these values may change accordingly.^6^

When we did intragroup comparisons of SpHb values in our study, no significant difference was observed between the blocked and non-blocked groups. Moreover, when we look at the comparison between the group of successful block and the group of failed block, no significant difference was reported.

Our study has some limitations. First, in our study, we could not achieve a successful block in five patients, and our objective parameter in these patients did not change significantly compared to baseline. Thus, the inability to produce significant increases in our objective parameter may be a reliable sign of block failure, but this should be confirmed in a larger-scale study with more cases of block patients. Secondly, we targeted patients scheduled to undergo any type of forearm surgery. The results of the study, which we have limited to one type of specific surgery, may not give an idea about how it will end in other specific surgeries. Thirdly, it is a common practice to sedate patients in peripheral nerve blocks. In our study, midazolam was given to the patients for sedation. Benzodiazepines can cause not only sedation but also amnesia and may impair the perception and identification of the senses, and mild sedation probably does not greatly impair the perception of the appearance of periarterial swelling caused by local anesthesia. Therefore, the applicability of our findings in sedation patients remains unclear.

When we compared the parameters among themselves, we found that, out of all the parameters, including StO_2_, THI, PI, SpHb and skin temperature, THI showed a significant difference between the blocked group and the control group, but there was no significant difference in terms of THI between the groups of successful block and failed block. In SpHb, no significant difference was neither reported between the block group and the control group nor between the groups of successful block and failed block. When we evaluated StO_2_, PI and body temperature parameters, we found that StO_2_ and PI presented with a significant difference between the groups of successful block and failed block at an earlier stage, compared to body temperature.

When we looked at the ROC curve for StO_2_, PI and body temperature parameters between the block and control groups, we found that StO_2_ was a more sensitive parameter than PI and body temperature.

### Conclusions

Of all the StO_2_, THI, PI, SpHb and skin temperature measurement parameters in infraclavicular plexus blocks, StO_2_, PI and skin temperature measurements are simple, objective, noninvasive techniques to evaluate the success or failure of the blocks. StO_2_ is the parameter with the highest specificity and sensitivity rate among these parameters, which start early and continue in the late period.

## Data Availability

Our data were obtained from some patients who underwent upper extremity surgery in the Orthopedics Service of Harran University Medical Faculty Hospital between 01/02/2019 and 01/12/2019, with the decision of the Harran University Ethics Committee. Data It can be obtained from Abdulhakim Şengel (ORCID: 0000-0003-0905-1018).

## ACKNOWLEDGEMENTS

We thank all included patients, all physicians, nurses, the surgical and anesthetic personnel for invaluable help. Further we thank Prof. Dr. Seval Kul from Gaziantep University Faculty of Medicine, Department of Biostatistics for guide our article statistically

